# Validation of an ICD-code-based case definition for psychotic illness across three health systems

**DOI:** 10.1101/2024.02.28.24303443

**Authors:** Anthony J. Deo, Victor M. Castro, Ashley Baker, Devon Carroll, Joseph Gonzalez-Heydrich, David C. Henderson, Daphne J. Holt, Kimberly Hook, Rakesh Karmacharya, Joshua L. Roffman, Emily M. Madsen, Eugene Song, William G. Adams, Luisa Camacho, Sarah Gasman, Jada S. Gibbs, Rebecca G. Fortgang, Chris J. Kennedy, Galina Lozinski, Daisy C. Perez, Marina Wilson, Ben Y. Reis, Jordan W. Smoller

**Affiliations:** Department of Psychiatry and Behavioral Sciences, Boston Children’s Hospital, Harvard Medical School, Boston, MA; Department of Psychiatry, Harvard Medical School, Boston, MA; Department of Psychiatry, Rutgers-Robert Wood Johnson Medical School, Piscataway, NJ; Rutgers University Behavioral Health Care, Piscataway, NJ; Research Information Science and Computing, Mass General Brigham, Somerville, MA; Ascend Integrative Medicine LLC / Massachusetts; University of Rhode Island, Providence, RI, USA; Tommy Fuss Center for Neuropsychiatric Disease Research, Boston Children’s Hospital, Harvard Medical School, Boston, MA; Early Psychosis Investigation Center, Boston Children’s Hospital, Harvard Medical School, Boston, MA; Boston Medical Center, Boston MA; Boston University Chobanian & Avedisian School of Medicine, Boston MA; Department of Psychiatry, Massachusetts General Hospital, Boston MA; Harvard T.H. Chan School of Public Health, Boston, MA; Center for Genomic Medicine, Massachusetts General Hospital, Boston, MA; Chemical Biology and Therapeutic Science Program, Broad Institute of MIT and Harvard, Cambridge, MA; Schizophrenia and Bipolar Disorder Program, McLean Hospital, Belmont, MA; Psychiatric & Neurodevelopmental Genetics Unit, Center for Genomic Medicine, Massachusetts General Hospital, Boston, MA, USA; Center for Precision Psychiatry, Department of Psychiatry, Massachusetts General Hospital, Boston, MA, USA; Rutgers New Jersey Medical School, Newark, New Jersey 07103; Department of Psychology, Harvard University, Cambridge, MA; Predictive Medicine Group, Harvard Medical School, Boston, MA; Computational Health Informatics Program, Boston Children’s Hospital, Boston, MA; Stanley Center for Psychiatric Research, Broad Institute, Cambridge, MA

**Keywords:** prediction, detection, schizophrenia, schizoaffective, bipolar

## Abstract

**Background and Hypothesis:** Early detection of psychosis is critical for improving outcomes. Algorithms to predict or detect psychosis using electronic health record (EHR) data depend on the validity of the case definitions used, typically based on diagnostic codes. Data on the validity of psychosis-related diagnostic codes is limited. We evaluated the positive predictive value (PPV) of International Classification of Diseases (ICD) codes for psychosis.

**Study Design:** Using EHRs at three health systems, ICD codes comprising primary psychotic disorders and mood disorders with psychosis were grouped into five higher-order groups. 1,133 records were sampled for chart review using the full EHR. PPVs (the probability of chart-confirmed psychosis given ICD psychosis codes) were calculated across multiple treatment settings.

**Study Results:** PPVs across all diagnostic groups and hospital systems exceeded 70%: Massachusetts General Brigham 0.72 [95% CI 0.68-0.77], Boston Children’s Hospital 0.80 [0.75-0.84], and Boston Medical Center 0.83 [0.79-0.86]. Schizoaffective disorder PPVs were consistently the highest across sites (0.80-0.92) and major depressive disorder with psychosis were the most variable (0.57-0.79). To determine if the first documented code captured first-episode psychosis (FEP), we excluded cases with prior chart evidence of a diagnosis of or treatment for a psychotic illness, yielding substantially lower PPVs (0.08–0.62).

**Conclusions:** We found that the first documented psychosis diagnostic code accurately captured true episodes of psychosis but was a poor index of FEP. These data have important implications for the development of risk prediction models designed to predict or detect undiagnosed psychosis.

## Introduction

Psychotic disorders typically have their onset during late adolescence or early adulthood, and are associated with high levels of lifelong disability and premature mortality.^1, 2^ Studies have demonstrated that decreasing the duration of untreated psychosis can improve functional outcomes.^3-5^ Substantial effort has been dedicated over the past several decades to developing approaches for early detection of those who are at risk for or in the early stages of psychosis, in order to facilitate early intervention.^6^

Electronic health records (EHRs) offer a large-scale resource of real-world health data and have been increasingly used in psychiatric research aimed at characterizing, detecting, or predicting important mental health outcomes.^7-12^ For example, a growing number of statistical and machine learning approaches have been applied to EHRs to identify or predict psychotic illness.^7-13^ Algorithms trained on EHR data typically rely on outcome labels in the form of International Classification of Diseases (ICD) diagnostic codes. The validity of these algorithms ultimately depends on how accurately these codes capture true psychosis, but studies evaluating this accuracy have been limited. In a study of first-episode psychosis using EHR and claims data from five health systems, Simon and colleagues^14^ conducted chart review validation for a subset of putative cases; there was substantial variability in PPVs of psychosis codes across healthcare settings ranging from 84% among patients age 15-29 in emergency settings to only 19% for those aged 30-59 diagnosed at general medical outpatient visits. Benson and colleagues^15^, using insurance claims data, found that only 23% of patients with a schizophrenia spectrum ICD code in a given year represented new onset cases based on up to 4 years of historical data.

In this study, as part of an effort to develop algorithms to detect undiagnosed psychosis, we evaluated the positive predictive value of initial ICD codes for psychotic illness across three independent healthcare systems. Using a systematic, expert clinician chart review of more than 1,100 records for patients age 15 – 35, we examined the following questions: 1) What is the probability that the first documented ICD diagnosis of psychotic illness accurately captures an episode of psychosis?; 2) How often is the first EHR psychosis code a new-onset (i.e. first-episode) of psychosis?; and, 3) Where available, how does the validity of ICD codes vary by treatment setting?

## Methods

The study was conducted at three health systems located in Boston, MA: Boston Children’s Hospital (BCH), Boston Medical Center (BMC), and Massachusetts General Brigham (MGB). BCH is a comprehensive center for pediatric health care, offering a complete range of health care services for children, adolescents, and young adults. BMC is an academic medical center and the largest safety net hospital in New England, caring for more uninsured patients than any other hospital in the region. MGB is the largest healthcare provider in Massachusetts, including Massachusetts General Hospital, Brigham and Women’s Hospital, McLean Hospital, and other major hospitals. EHR data were collected from each site with a waiver of consent obtained from the institutional review board at MGB which served as the single IRB. Data from MGB is extracted from the Research Patient Data Repository (RPDR), containing EHR data for over 3 million patients in an i2b2 common data model. The dataset for this study includes demographics, diagnosis, medications, procedures, and lab tests from 1998 to 2022 for patients with at least 3 visits greater than 90 days apart. Data from BMC is extracted from the Research Data Warehouse, containing EHR data for 2.5 million patients in an i2b2 common data model. Data from BCH is extracted from the BCH Enterprise Data Warehouse (EDW), containing data on over 1.8M patients in an i2b2 common data model. The dataset for this study includes demographics, diagnosis, medications, procedures, and lab tests from 1990 to 2022 for patients with at least 3 visits greater than 90 days apart.

Records were identified for chart review if at least 2 psychosis associated diagnostic codes were present, the first of which was a code associated with a nonchronic form of psychosis (**see Supplemental Methods, Supplemental Tables 1,2,3, and Supplemental Figure 1**). The first documented code was considered the “index code”. Diagnostic codes associated with substance- or general medical condition (GMC)-induced psychotic disorders were neither inclusionary nor exclusionary criteria for this study. Eligible records were restricted to individuals between the ages of 15 and 35 years old at the time of the first psychosis diagnostic code and whose first code was on or after January 1, 2000 (**Supplemental Figure 1**). As in prior research^14, 15^, we focused on the age range of 15 – 35, as this corresponds to the typical age of onset for psychotic illness. To ensure adequate clinical information, we also required at least two encounters 2 years or more prior to the index code date, at least one of which occurred within 10 years prior to the index code date.

All records were categorized into one of the following five code groups according to their index code (**Supplemental Table 1** for a list of ICD codes included in each grouping)): schizophrenia unspecified state (ICD 9: 295, 295[.0, .00, .1, .10, .2, .20, .3, .30, .4, .40, .5, .50, .8, .80, .9, .90, .91, .92, .93, .94], ICD10: F20, F20[.0, .1, .2, .3, .5, .8, .81, .89, .9]), schizoaffective disorder (ICD 9: 295.7, 295.70, ICD10: F25, F25[.0, .1, .8, .9]), other unspecified psychosis and/or psychotic disorders (ICD 9: 297, 297[.0, .1, .2, .3, .8, .9] 298[.1, .3, .4, .8, .9], ICD10: F22, F23, F28, F29, DRG:430, APDRG:430), major depressive disorder with psychosis (ICD 9: 296.24, F296.34, ICD 10: F32.3, F33.3) and bipolar disorder with psychosis (ICD 9: 296[.04, .14, .44, .54, .64], ICD 10: F30.2, F31[.2, .5, .64]). At each of the three participating health systems, 100 cases from each code group were randomly sampled for chart review (with the exception of BCH where most code group’s available sample size was less than 100 (ranging from 20 to 100 records per group)). Case sampling included all encounters at the individual institutions, including all medical and psychiatric inpatient, outpatient and emergency department settings. We then excluded records where no clinical notes were available within 60 days before or after the date of the first diagnostic code as well as notes from McLean hospital prior to 2018 (before records were available for research use).

At each participating site, two clinicians (psychiatrist, psychologist, or psychiatric advanced practice nurse practitioner) with expertise in diagnosing and treating psychotic disorders independently reviewed each patient’s full-text EHR including all available notes from all settings. If there was a discrepancy in the chart review diagnosis, a third senior psychiatrist served as a tie-breaker. A series of pilot chart reviews were completed to develop and refine the case definition and develop the structured chart review tool (**Supplemental Figure 2**). All reviewers completed 2-4 hours of training and interrater calibration prior to chart review.

The initial step in the review was to determine if there was evidence of psychosis in the chart within a 60 day window around the first (index) diagnostic code (**Figure 1**). This evidence of psychosis could consist of: an EHR note with a formal diagnosis of psychosis or recorded positive symptoms of psychosis (delusions, hallucinations, disorganized behavior) (see **Supplement Figure 2** for chart review instrument). Note that this determination was made without consideration of the presence or absence of antipsychotic medication because the indications for antipsychotic use are not specific to psychosis (e.g. antidepressant augmentation, mood stabilization). Records were not classified as a case if the chart reviewers determined that the symptoms of psychosis occurred only in the presence of a substance or GMC and resolved in the absence of a substance or GMC (**Figure 1**).

**Figure 1:**
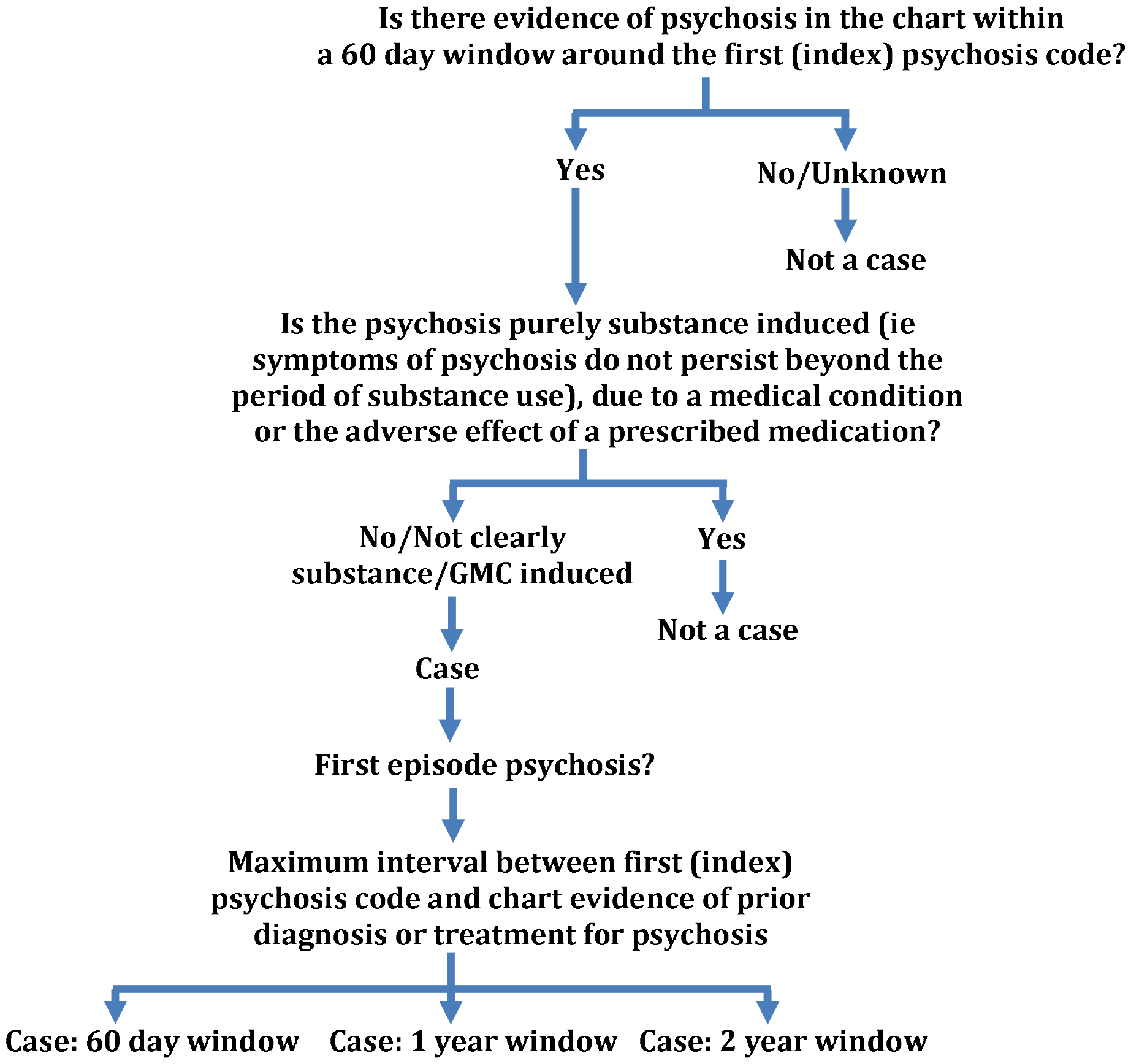
Each chart was examined during the window of 60 days before the first psychosis (index) diagnostic code to 60 days after. If there was evidence of psychosis in the chart in this time window and it was not purely substance or GMC induced psychosis, the record was defined as a case. The chart was then examined to identify the maximum interval between the first (index) psychosis code and chart evidence of prior diagnosis or treatment for psychosis. These time windows included 60 days, 1 year or 2 years before the index code.

For each of the 5 case groupings, the validity of the case definitions was calculated as positive predictive values (PPVs) and their 95% confidence intervals—that is, the probability of being a case of psychotic illness by chart review given the ICD code group label. We also examined whether the index code represented an FEP by applying 3 different time windows for excluding prior evidence of psychosis. We first required that there be no evidence in narrative notes of a prior formal diagnosis of a psychotic disorder or specific treatment of psychosis more than 60 days prior to the index code. Note that records were considered a case if they had prior symptoms of psychosis without a formal diagnosis of a psychotic illness or there was documented use of an antipsychotic for indications other than psychosis when defining case status for the temporal windows. Because an initial analysis using these criteria yielded PPVs < .70, we next relaxed these duration criteria to exclude psychosis 1 year or 2 years prior to the index code, consistent with broader criteria for FEP that have previously been suggested. (**Figure 1 and Supplemental Figure 3**). Cases that did not meet criteria based on the 60-day temporal window were re-reviewed with these relaxed temporal windows for achieving case status. One reviewer at each site examined notes from the prior chart review and, if needed EHR charts, to make this determination (See **Supplemental Figure 4** for Chart Review Instrument for this portion of the study). In an exploratory analysis, we examined PPVs according to the treatment setting (emergency department, outpatient, or inpatient) where the index code was given. Each setting included all medical and psychiatric encounters, only BMC outpatient visits were separated into psychiatric and non-psychiatric visits. Setting-specific data was available for subsets of records at all sites (MGB (93%), BCH (63%) and BMC (95%)).

## Results

### Does the first ICD code for psychotic illness code coincide with a documented episode of psychosis?

We first conducted chart review validation for the presence of psychotic illness within 60-days around the index psychosis code for each code group. Among all institutions there were 1,133 patient records which met the selection criteria for chart review. (MGB 356, BCH 284, BMC 493). As shown in Table 1, PPVs across the 5 code groups exceeded 70% across institutions: MGB 0.72 [95% CI 0.68-0.77], BCH 0.80 [0.75-0.84], BMC 0.83 [CI 0.79-0.86], indicating generally high concordance between ICD codes and expert clinician judgment. Within each health system, the schizoaffective disorder code group had the highest PPVs: MGB 0.80 (95% CI: 0.69-0.90), BCH 0.90 (0.80-1.01), BMC 0.92 (0.87-0.97). The lowest PPV was observed at MGB for major depressive disorder with psychosis [0.57(0.45-0.69)], though PPVs were higher for this group at BCH 0.73 (0.63 – 0.83) and BMC 0.79 (0.71 – 0.87). Other code groups showed some variability by site in terms of PPV, though most were in the range of 0.69 – 0.91.

**Table 1.** Positive predictive values (PPVs) by diagnostic code group across three healthcare systems.

|  | Psychosis<br>present |  | 60 day |  | 1 year |  | 2 years |  |  |
| --- | --- | --- | --- | --- | --- | --- | --- | --- | --- |
| Code Group | PPV (N) | 95% CI | PPV (N) | 95% CI | PPV (N) | 95% CI | PPV (N) | 95% CI | Total N |
| Massachusetts General Brigham (MGB) |  |  |  |  |  |  |  |  |  |
| Schizophrenia Unspecified State | 0.69 (53) | 0.58 - 0.79 | 0.14 (11) | 0.06 - 0.22 | 0.17 (13) | 0.09 - 0.25 | 0.21 (16) | 0.12 - 0.30 | 77 |
| Schizoaffective Disorder | 0.80 (47) | 0.69 - 0.90 | 0.22 (13) | 0.11 - 0.33 | 0.24 (14) | 0.13 - 0.35 | 0.25 (15) | 0.14 - 0.37 | 59 |
| Other Unspecified Psychosis | 0.75 (69) | 0.66 - 0.84 | 0.51 (47) | 0.41 - 0.61 | 0.58 (53) | 0.48 - 0.68 | 0.59 (54) | 0.49 - 0.69 | 92 |
| Major Depressive Disorder with Psychosis | 0.57 (37) | 0.45 - 0.69 | 0.48 (31) | 0.36 - 0.60 | 0.51 (33) | 0.39 - 0.63 | 0.51 (33) | 0.39 - 0.63 | 65 |
| Bipolar Disorder with Psychosis | 0.81 (51) | 0.71 - 0.91 | 0.60 (38) | 0.48 - 0.72 | 0.62 (39) | 0.50 - 0.74 | 0.62 (39) | 0.50 - 0.74 | 63 |
| Overall | 0.72 (257) | 0.68 - 0.77 | 0.39 (140) | 0.34 - 0.44 | 0.43 (152) | 0.38 - 0.48 | 0.44 (157) | 0.39 - 0.49 | 356 |
| Boston Children's Hospital (BCH) |  |  |  |  |  |  |  |  |  |
| Schizophrenia Unspecified State | 0.88 (59) | 0.80 - 0.96 | 0.25 (17) | 0.15 - 0.36 | 0.40 (27) | 0.29 - 0.52 | 0.51 (34) | 0.39 - 0.63 | 67 |
| Schizoaffective Disorder | 0.90 (28) | 0.80 - 1.01 | 0.10 (3) | -0.01 - 0.20 | 0.23 (7) | 0.08 - 0.37 | 0.26 (8) | 0.10 - 0.41 | 31 |
| Other Unspecified Psychosis | 0.73 (68) | 0.64 - 0.82 | 0.46 (43) | 0.36 - 0.56 | 0.49 (46) | 0.39 - 0.60 | 0.55 (51) | 0.45 - 0.65 | 93 |
| Major Depressive Disorder with Psychosis | 0.73 (55) | 0.63 - 0.83 | 0.44 (33) | 0.33 - 0.55 | 0.53 (40) | 0.42 - 0.65 | 0.57 (43) | 0.46 - 0.69 | 75 |
| Bipolar Disorder with Psychosis | 0.89 (16) | 0.74 - 1.03 | 0.39 (7) | 0.16 - 0.61 | 0.50 (9) | 0.27 - 0.73 | 0.50 (9) | 0.27 - 0.73 | 18 |
| Overall | 0.80 (226) | 0.75 - 0.84 | 0.36 (103) | 0.31 - 0.42 | 0.45 (129) | 0.40 - 0.51 | 0.51 (145) | 0.45 - 0.57 | 284 |
| Boston Medical Center (BMC) |  |  |  |  |  |  |  |  |  |
| Schizophrenia Unspecified State | 0.91 (89) | 0.85 - 0.97 | 0.16 (16) | 0.09 - 0.24 | 0.20 (20) | 0.12 - 0.28 | 0.31 (30) | 0.21 - 0.4 | 98 |
| Schizoaffective Disorder | 0.92 (93) | 0.87 - 0.97 | 0.08 (8) | 0.03 - 0.13 | 0.10 (10) | 0.04 - 0.16 | 0.15 (15) | 0.08 - 0.22 | 101 |
| Other Unspecified Psychosis | 0.75 (83) | 0.67 - 0.83 | 0.27 (30) | 0.19 - 0.36 | 0.29 (32) | 0.21 - 0.38 | 0.29 (32) | 0.21 - 0.38 | 110 |
| Major Depressive Disorder with Psychosis | 0.79 (80) | 0.71 - 0.87 | 0.35 (35) | 0.25 - 0.44 | 0.42 (42) | 0.32 - 0.51 | 0.47 (47) | 0.37 - 0.56 | 101 |
| Bipolar Disorder with Psychosis | 0.75 (62) | 0.65 - 0.84 | 0.17 (14) | 0.09 - 0.25 | 0.20 (17) | 0.12 - 0.29 | 0.25 (21) | 0.16 - 0.35 | 83 |
| Overall | 0.83 (407) | 0.79 - 0.86 | 0.21 (103) | 0.17 - 0.24 | 0.25 (121) | 0.21 - 0.28 | 0.29 (145) | 0.25 - 0.33 | 493 |

### How often does the first psychosis ICD code represent a new-onset of psychotic illness (i.e. FEP)?

After determining that the index psychosis ICD codes were generally valid indicators of documented psychotic illness, we evaluated whether the index code captures FEP. As described earlier, we did this by evaluating 3 temporal windows for defining FEP, requiring that there be no formal diagnosis of or specific treatment for a psychotic illness prior to a) 60 days; b) 1 year; or c) 2 years before the index code. As shown in Table 1, we observed low PPVs for FEP (range 0.08 – 0.62), with the lowest values seen in the 60 day window (range 0.08 – 0.60). There was minimal improvement in PPVs when expanding to the 1 and 2 year windows (PPVs ranging from 0.10 to 0.62 across all sites and code groups). Substantial variability in PPVs were observed by code group and site, but perhaps unsurprisingly, the highest values were observed at BCH which serves a child and adolescent patient population. Nevertheless, these findings suggest that the first psychosis code documented in a health system is unlikely to represent FEP.

### How does the validity of ICD codes vary by treatment setting?

As noted in the Methods section, we distinguished three settings (ED, inpatient, and outpatient) according to the visit at which the index code was given. At BMC, we were further able to distinguish outpatient psychiatric vs non psychiatric visits. Of note there was a difference in the distribution of setting across sites. At BMC 45% of records where the setting was known originated from the emergency department whereas this represented only 8% of the records at MGB and 21% of records at BCH (**Table 2**).

**Table 2:** Positive predictive values (PPVs) for confirmed psychosis across settings.

|  | Setting | PPV (N) | 95% CI | Total N |
| --- | --- | --- | --- | --- |
| **Massachusetts General Brigham (MGB)** |  |  |  |  |
|  | Emergency | 0.80 (20) | 0.64 - 0.96 | 25 |
|  | Inpatient | 0.70 (80) | 0.61 - 0.78 | 115 |
|  | Outpatient | 0.73 (138) | 0.66 - 0.79 | 190 |
| **Boston Children's Hospital (BCH)** |  |  |  |  |
|  | Emergency | 0.82 (31) | 0.69 - 0.94 | 38 |
|  | Inpatient | 0.91 (29) | 0.81 - 1.01 | 32 |
|  | Outpatient | 0.80 (88) | 0.73 - 0.87 | 110 |
| **Boston Medical Center (BMC)** |  |  |  |  |
|  | Emergency | 0.89 (188) | 0.84 - 0.93 | 212 |
|  | Inpatient | 0.70 (21) | 0.54 - 0.86 | 30 |
|  | Outpatient (nonpsych) | 0.77 (101) | 0.70 - 0.84 | 131 |
|  | Outpatient (psych) | 0.84 (80) | 0.77 - 0.92 | 95 |

With these limitations in mind, we observed generally comparable performance across settings for capturing psychotic illness overall (as opposed to FEP). For the ED setting, PPVs ranged from 0.80 (0.64-0.96) at MGB to 0.89 (0.84-0.93) at BMC. For index codes assigned at inpatient admissions, PPVs were 0.70 at both MGB and BMC but 0.91 (0.81 – 1.01) at BCH. Outpatient codes had similar performance across sites ranging from 0.73 – 0.84. At BMC, codes given at psychiatric outpatient visits (PPV = 0.84) slightly outperformed those given at non-psychiatric visits (PPV = 0.77). Across the three health systems, differences among PPVs by visit type had largely overlapping 95% CI, suggesting that point estimates were not meaningfully different.

## Discussion

Electronic health records have become an increasingly common resource for psychiatric research, including the development of diagnostic and predictive algorithms for mental health outcomes based on ICD codes. The validity of such research depends importantly on the degree to which these diagnostic codes accurately capture the target phenotype. As a prelude to developing algorithms to detect undiagnosed psychotic illness, we conducted a systematic validation of a range of psychosis-related ICD codes using expert clinician chart review of 1,133 patient records across three large healthcare systems.

Several findings from this effort are noteworthy. First, we found that an initial psychosis code was relatively accurate in capturing psychotic presentations across all hospital systems. Overall, when an index psychosis code was given, the probability that it was accompanied by chart documentation of psychotic illness ranged from 72% to 83%. We grouped ICD codes into 5 higher order categories of affective or nonaffective psychosis. With the exception of major depressive disorder with psychosis at one hospital system, PPVs for all code groups were high (range 0.69 – 0.92). These findings suggest that ICD codes are appropriate for use in EHR-based research involving the prediction or detection of psychosis. Second, we found that the accuracy of index codes for capturing psychotic illness was largely comparable across ED, inpatient, and outpatient settings, with PPVs ranging from a low of 0.70 to a high of 0.91 for the three healthcare systems. Third, however, we found that overall the first psychosis code documented in the EHR was unlikely to represent a first-episode psychosis (FEP) when this was defined as having no evidence in a patient’s notes of prior formal diagnosis of or specific treatment for a psychotic illness either 60 days (PPVs = 0.21 – 0.39), 1 year (PPVs = 0.25 – 0.45) or 2 years (PPVs = 0.29 – 0.51) prior to the index code. It is important to note, however, that each of the three healthcare systems is an “open” (rather than fully integrated) system. That is, not all healthcare encounters would be observed if patients received care from outside facilities; this may partially or fully account for the mismatch between index codes and FEP. Nevertheless, our results are broadly consistent with findings from a study^15^ that used claims data to identify index cases of schizophrenia spectrum disorders in a given year (2016) among Massachusetts patients age 15-35. The authors found that historical data up to 4 years prior to the index ICD code revealed that 77% had evidence of a prior psychotic disorder diagnosis. These results accord with our finding that an apparent index ICD diagnosis of psychotic illness may not represent true FEP.

To our knowledge, only one prior study has systematically validated the accuracy of ICD psychosis codes. In a population-based study of FEP across 5 healthcare systems in the Kaiser Permanente network, Simon and colleagues^14^ reviewed 1337 records of patients age 15 – 59 years at the time of a first-occurring affective or non affective ICD psychotic illness code. This study included diagnostic codes derived from both EHRs and insurance claims, reducing the likelihood of unobserved psychotic illness. In addition, Simon et al. focused specifically on the incidence of FEP (defined as no evidence of a prior diagnosis of or treatment for a psychotic disorder more than 60 days prior to the index code), and charts were selected by random sampling stratified by age (above or below 30 years) and setting (mental health inpatient/ED, mental health outpatient, general medical outpatient). Compared to our random sampling stratified by 5 diagnostic code groups, this sampling scheme resulted in somewhat different distributions of ICD codes among the charts selected for review. Given these considerations, the PPVs for FEP reported by Simon et al. are not directly comparable to those we observed, though they encompassed a wide range of values—from 84% for patients age 15-29 diagnosed in inpatient psychiatric settings to 19% for patients age 30-59 diagnosed in primary care settings.

Our study included notable strengths including the incorporation of data from three large, independent healthcare systems encompassing both adult and pediatric settings. In addition, we used a systematic, structured chart review process conducted by expert clinicians. On the other hand, our results should be interpreted in light of several limitations. First, our sampling strategy for index cases included a data floor requiring at least one psychosis code following the index code (to enrich for true cases) and at least two documented encounters 2 years or more prior to the index code date (to enrich for patients with historical data). As such, our results may not apply to index codes selected without these restrictions. Second, as noted earlier, our estimates of the accuracy of index codes for identifying FEP may be inflated if first episodes were not documented in the EHRs of the participating health systems. Third, although we reviewed a large number of records (1,133), we did not have sufficient data to estimate PPVs separately for each ICD code included in the 5 code groups. In addition, although we attempted to review 100 records for each code group (i.e., 1500 in total), some code groups at each site did not have sufficient records meeting our selection criteria.

In summary, in a systematic chart review of longitudinal health records from 1,133 patients across three healthcare systems, we found that first documented ICD codes for psychotic illness are likely to reflect true cases with reasonable accuracy (PPVs 0.72 – 0.83). In contrast, they are poor indicators of FEP, even allowing for prior psychotic symptoms or treatment up to two years earlier. This study was undertaken to validate outcome labels for training machine learning algorithms that could detect undiagnosed or incipient psychosis (by estimating the probability of psychosis among patients lacking a documented diagnosis). Our results suggest that the selection of cases (and non-cases) according to the selection strategy used here is well-justified.

## Data Availability

Protected Health Information restrictions for individual-level apply to the availability of the clinical data here, which were IRB approved for use only in the current study. As a result, this dataset is not publicly available.

## Acknowledgments

This research was supported by NIMH R01 MH116042 (Smoller/Reis). Dr. Deo is supported by the National Center for Advancing Translational Sciences of the National Institutes of Health under Award Number KL2TR003018. Dr. Fortgang is supported by the National Institute of Mental Health (K23MH132766). The content is solely the responsibility of the authors and does not necessarily represent the official views of the National Institutes of Health.

## Disclosures

Dr. Smoller is a member of the Scientific Advisory Board of Sensorium Therapeutics (with equity), and has received grant support from Biogen, Inc. He is PI of a collaborative study of the genetics of depression and bipolar disorder sponsored by 23andMe for which 23andMe provides analysis time as in-kind support but no payments. Dr. Henderson is a member of the Data Safety Monitoring Board for Sunovion Pharmaceutical. Dr. Gonzalez-Heydrich is the founding head of the scientific advisory board of and has equity in Mightier/Neuromotion Labs creators of emotional regulation training video games. He has been a consultant to Sunovion, Neurocrine, and Alkermes pharmaceutical companies. All remaining authors have no financial disclosures.

## Notes

### Author Declarations

The study was conducted at three health systems located in Boston, MA: Boston Childrens Hospital (BCH), Boston Medical Center (BMC), and Massachusetts General Brigham (MGB). BCH is a comprehensive center for pediatric health care, offering a complete range of health care services for children, adolescents, and young adults. BMC is an academic medical center and the largest safety net hospital in New England, caring for more uninsured patients than any other hospital in the region. MGB is the largest healthcare provider in Massachusetts, including Massachusetts General Hospital, Brigham and Womens Hospital, McLean Hospital, and other major hospitals. EHR data were collected from each site with a waiver of consent obtained from the institutional review board at MGB which served as the single IRB.

