## Supplemental Figure 1 for "Validation of an ICD-code-based case definition for psychotic illness across three health systems"

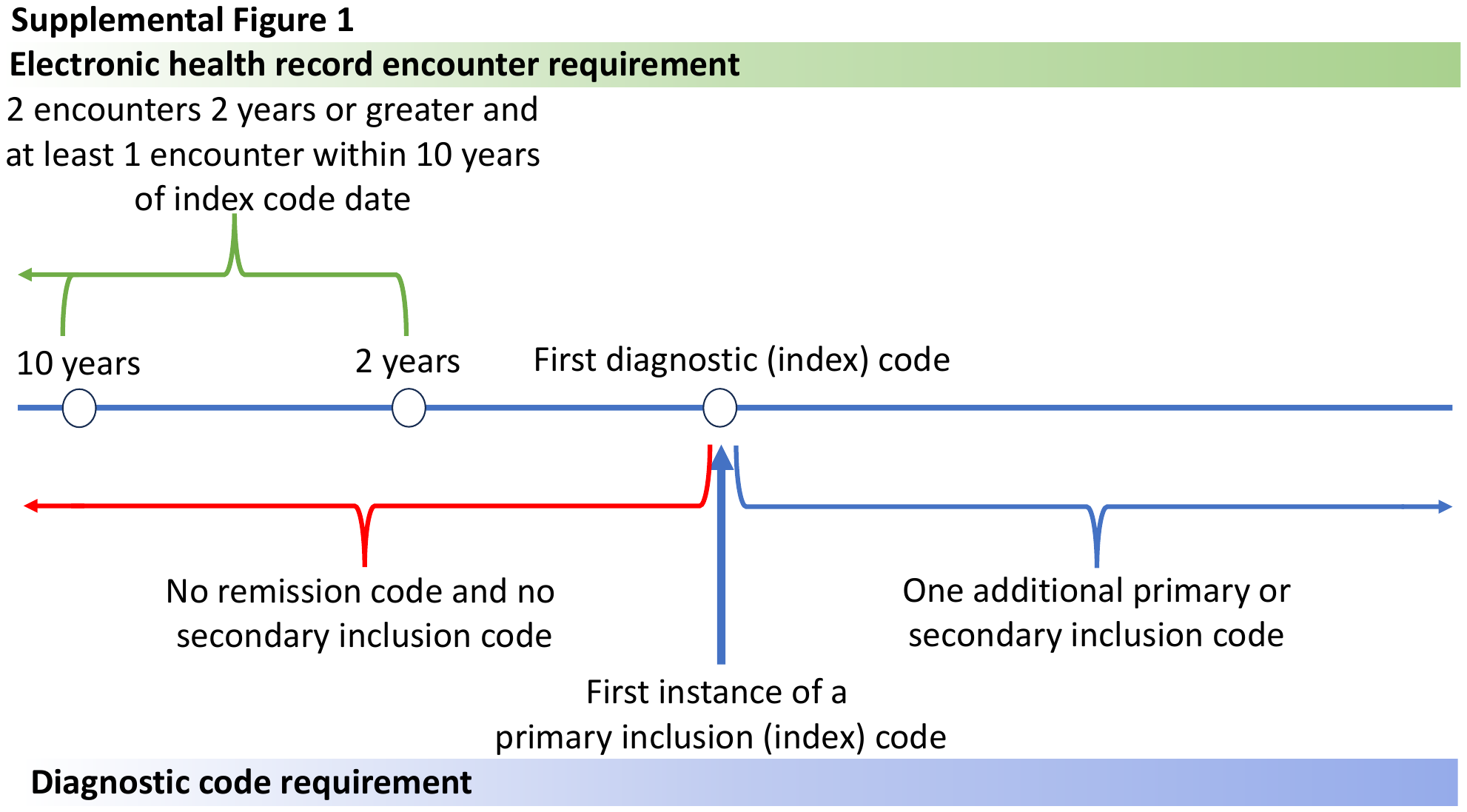

**Supplemental Figure 1:** Criteria used to identify charts for review. Bottom is a representation of the chronological order of diagnostic codes used for inclusion. Charts must include a primary inclusion (index) code at the index code date, with no secondary inclusion code or remission code prior to the index code date. Charts must also include at least one other primary or secondary inclusion code following the index code date. Top are the electronic health record encounter requirements for inclusion which included at least 2 encounters 2 years or more before the index code date and at least 1 encounter within 10 years before the index code date
