## Supplementary figures and images for "Validation of an ICD-code-based case definition for psychotic illness across three health systems"

### Supplemental Figure 2

**Supplemental Figure 2: Chart review data extraction instrument**
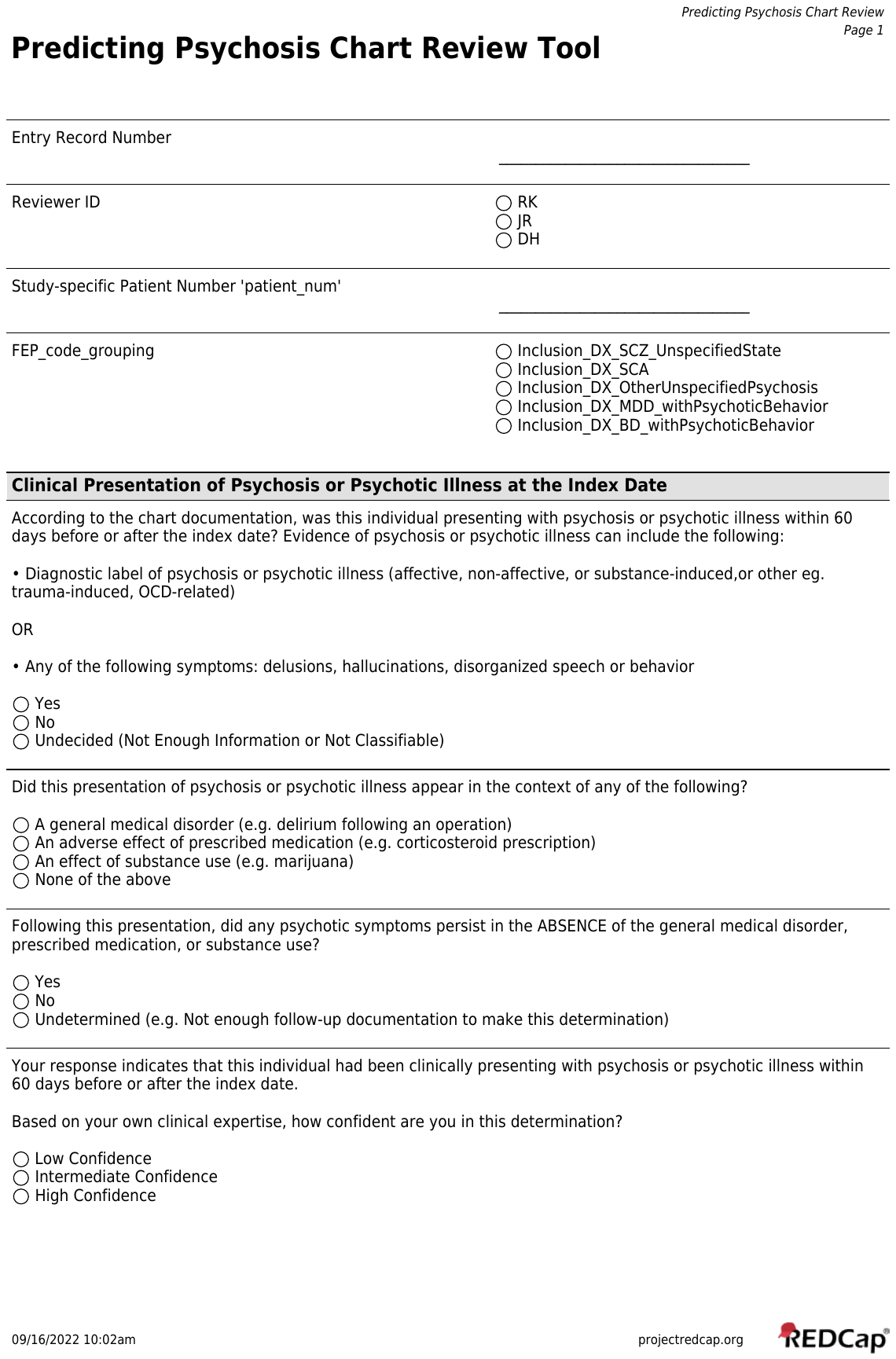

### Supplemental Table 1

#### **Supplemental Table 1: Primary Inclusion Codes**

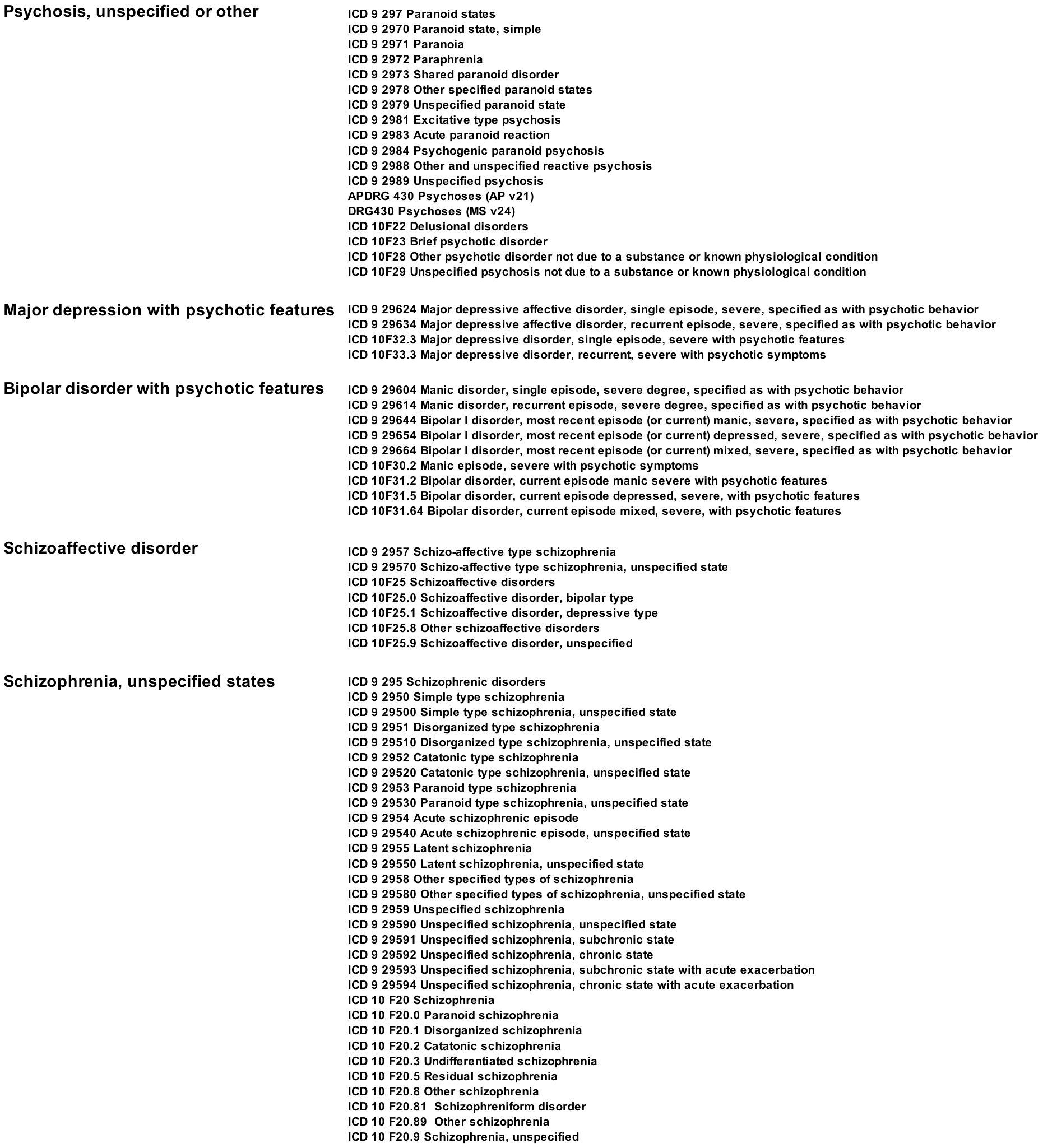

### Supplemental Table 2

#### **Supplemental Table 2: Remission Codes**

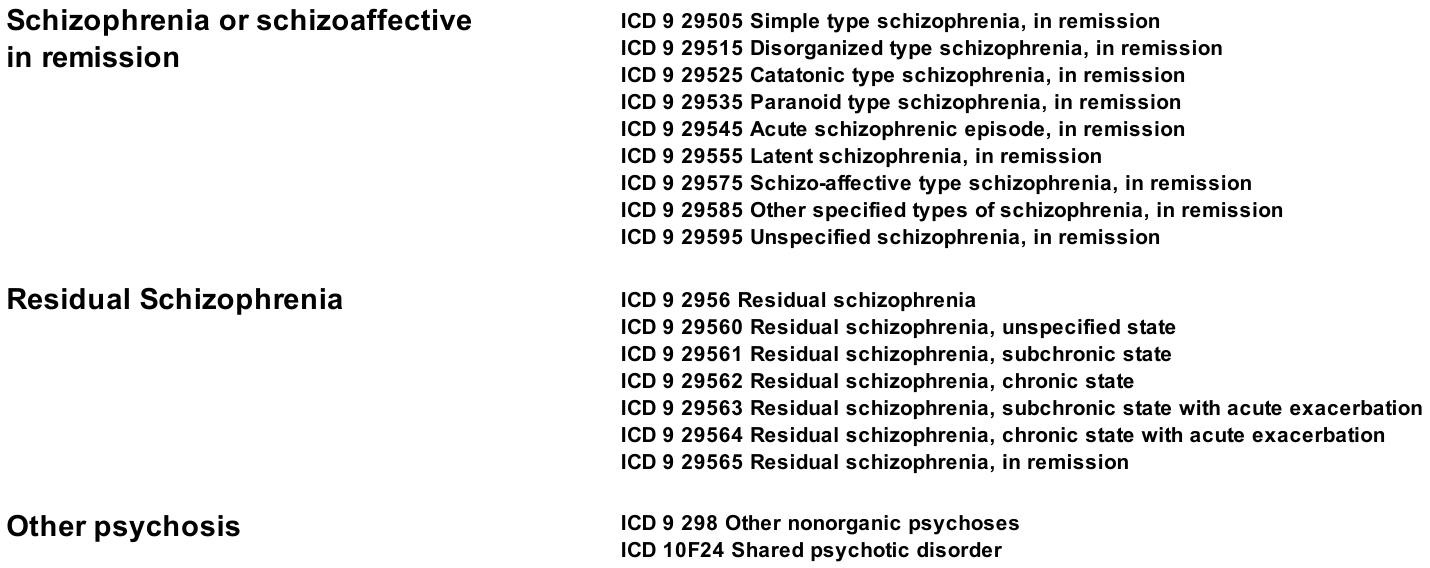

### Supplemental Table 3

#### **Supplemental Table 3: Secondary Inclusion Codes**

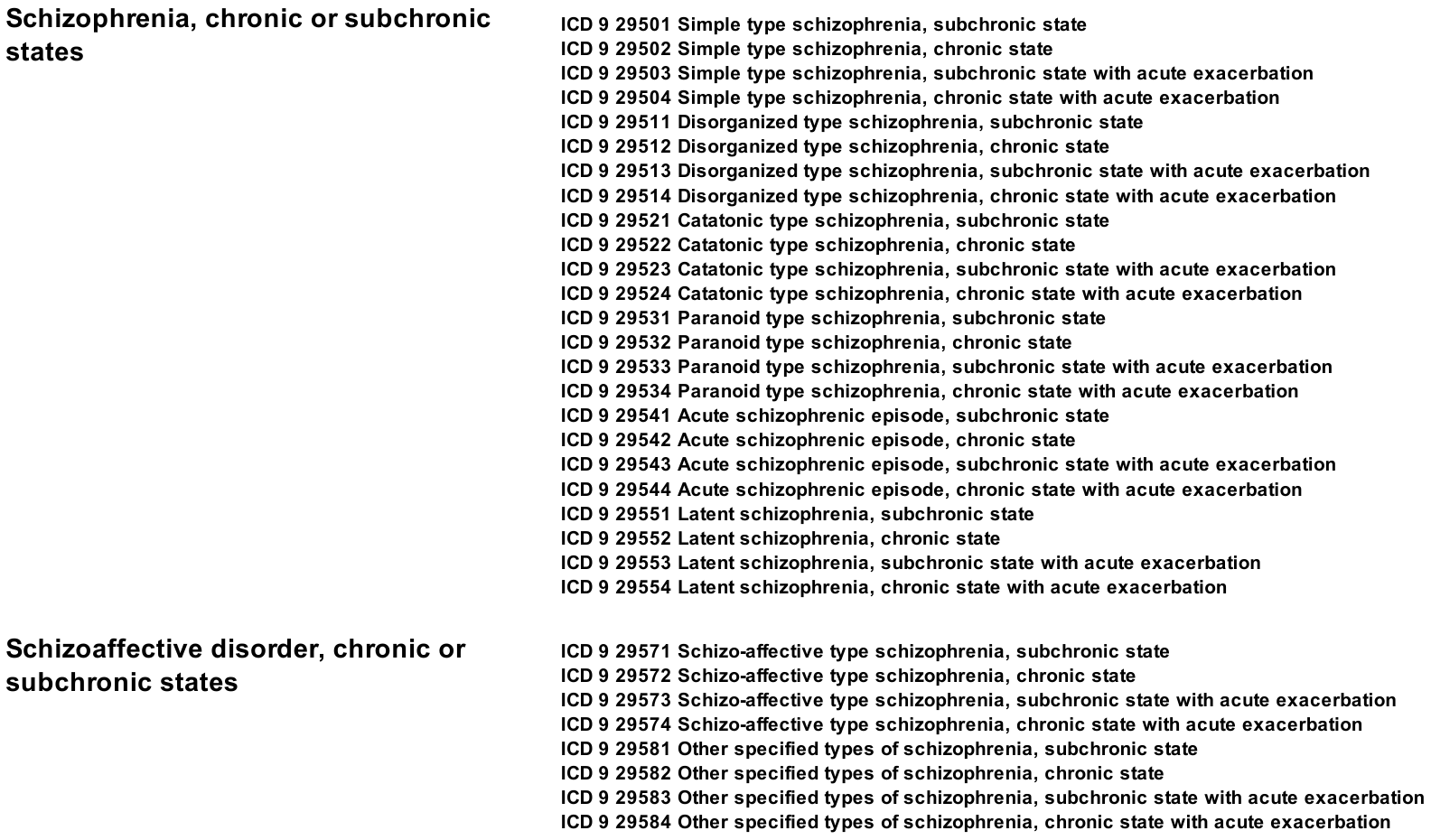
