## Supplemental Figure 4 for "Validation of an ICD-code-based case definition for psychotic illness across three health systems"

**Supplemental Figure 4: Chart review data extraction instrument used with expanded time windows**
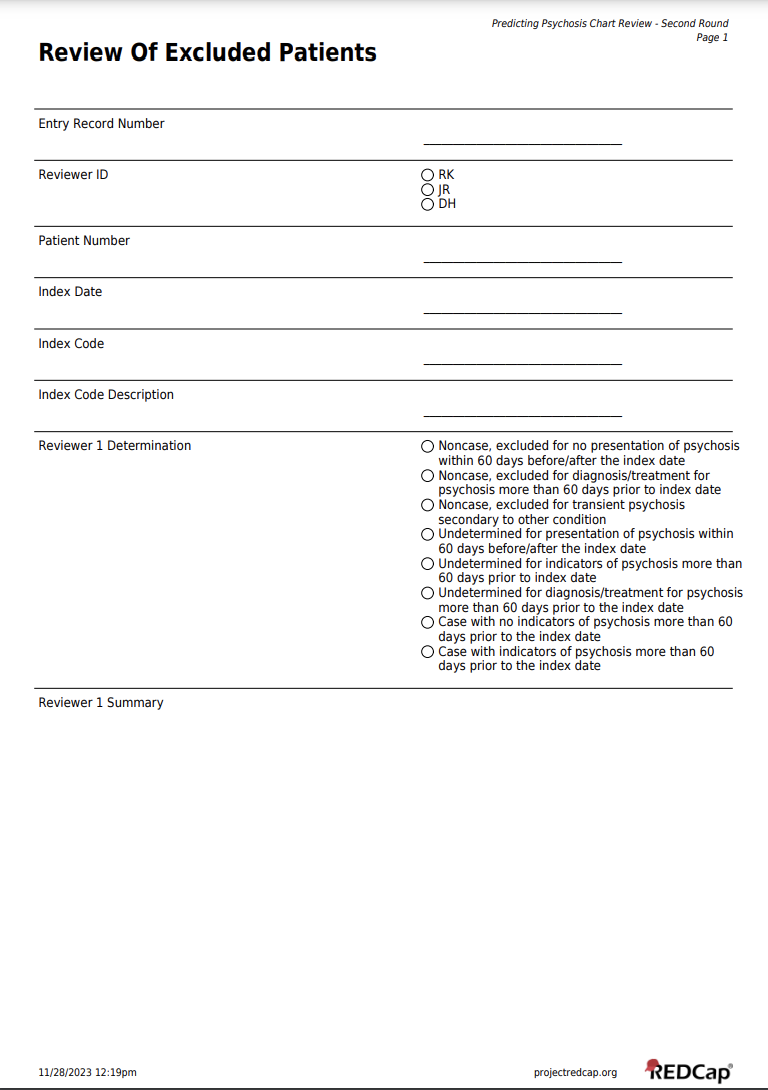

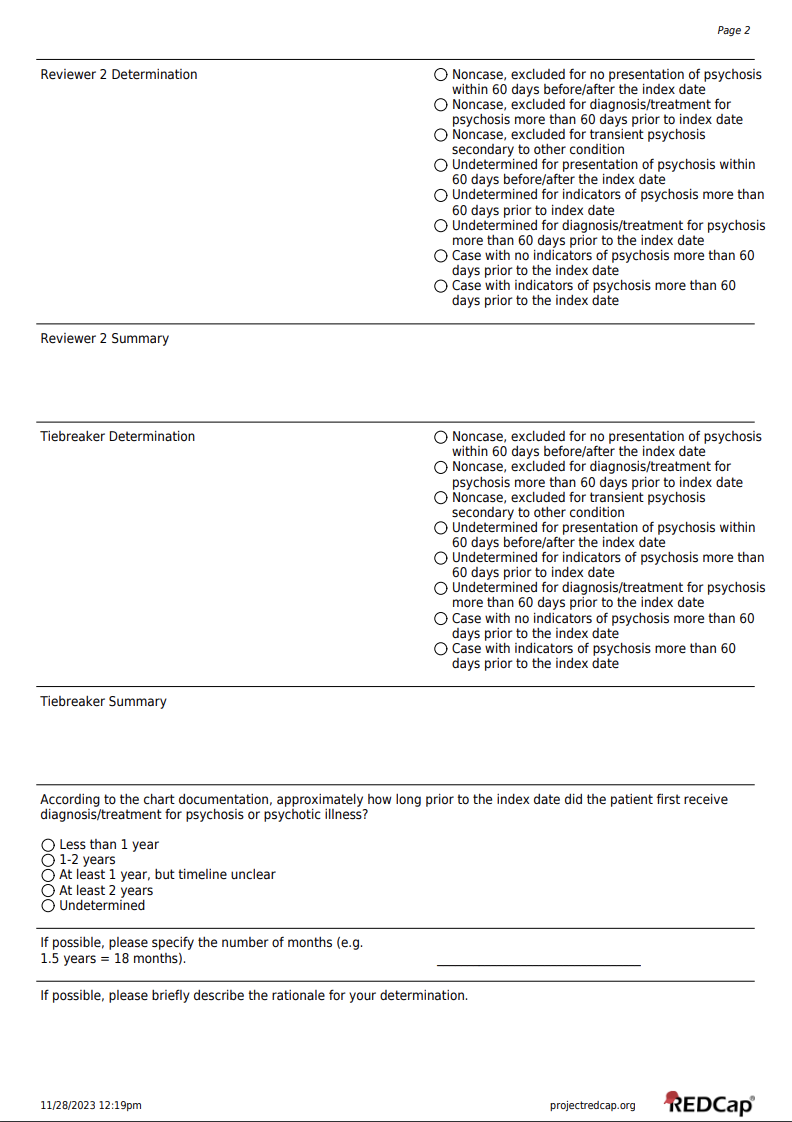
