## Supplemental Methods for "Validation of an ICD-code-based case definition for psychotic illness across three health systems"

ICD Diagnostic Codes used for inclusion and exclusion: Psychosis-associated diagnostic codes were partitioned into 3 groups and utilized to identify records for chart review as follows (**Supplemental Figure 1**). **Primary inclusion codes** included codes for primary psychotic disorders and affective disorders with psychosis without a chronic or subchronic designation (**Supplemental Table 1**). **Remission codes** are diagnostic codes indicating the underlying psychotic disorder is in remission or defined as residual (**Supplemental Table 2**). **Secondary inclusion codes** are associated with a chronic or subchronic psychosis diagnosis (**Supplemental Table 3**). Diagnostic codes associated with substance- or general medical condition (GMC)-induced psychotic disorders were neither inclusionary nor exclusionary criteria for this study.

Identification of records for potential chart review: Records were initially identified via the presence of an ICD diagnostic code associated with non-chronic forms of psychosis (i.e. primary inclusion codes). At each participating site, individual patient records were selected for chart review according to the following rule which required: 1) identifying the first instance of a primary inclusion (index) code; 2) the presence of a second code (primary or secondary inclusion code) following the index code; thus the second code allows for either non-chronic or chronic psychosis; 3) no secondary inclusion code or remission code before the index code date.
