## Supplemental Figure 3 for "Validation of an ICD-code-based case definition for psychotic illness across three health systems"

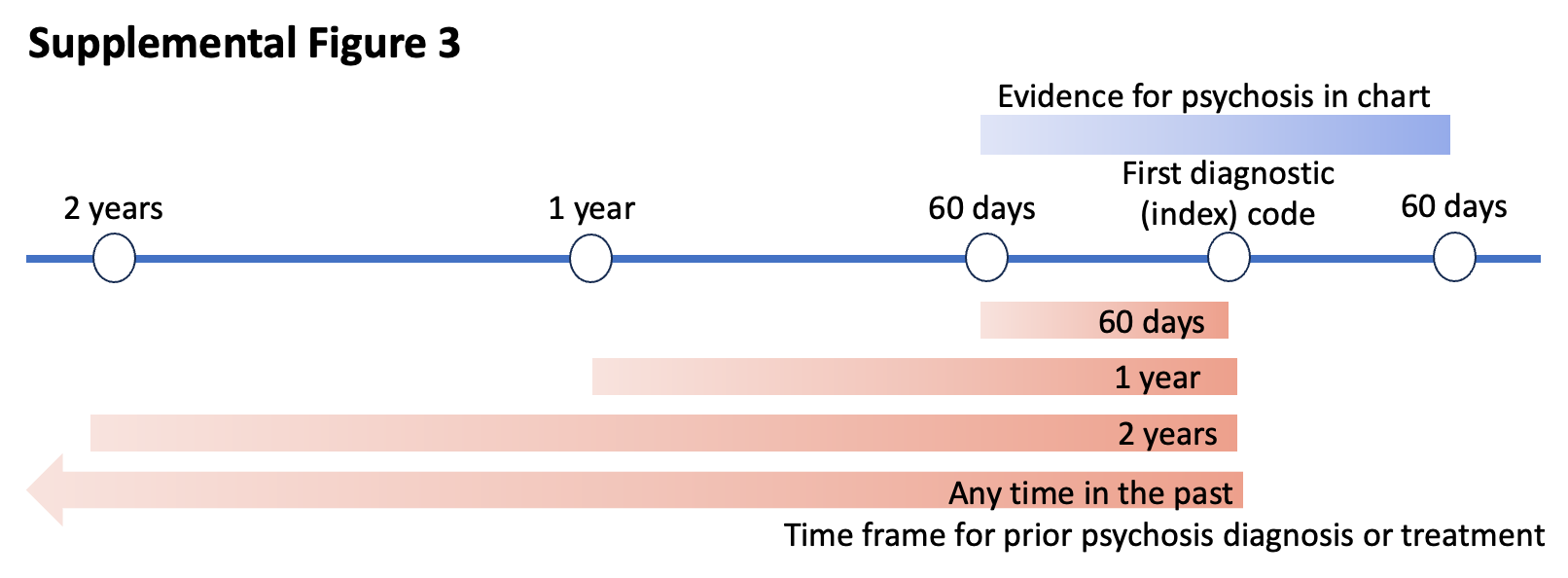

**Supplemental Figure 3:** Each chart was examined in a window +/- 60 days from the index diagnosis date (top in blue). If there was evidence of psychosis present in the chart in this time window, the chart was then examined to identify the time window for which there was a first reported diagnosis of or formal treatment for psychosis in the past (bottom in red). These time windows included 60 days, 1 year or 2 years before the first (index) diagnostic code or at any time in the past.
